# The role of *CYP2C19* gene polymorphisms on antiplatelet activity of clopidogrel among Arabs: systematic review and meta-analysis

**DOI:** 10.1101/2022.02.01.22270244

**Authors:** Abdullah N. Alkattan, Nashwa M. Radwan, Nagla E. Mahmoud, Amjad F. Alfaleh, Amal Alfaifi, Khaled I. Alabdulkareem

**Affiliations:** Department of Research and Development, General Directorate of School Health, Ministry of Health, Riyadh, Saudi Arabia; Department of Public Health and Community Medicine, Faculty of Medicine, Tanta University, Tanta, Egypt; Department of Research, Assisting Deputyship for Primary Health Care, Ministry of Health, Riyadh, Saudi Arabia; Department of Family Medicine, College of Medicine, Al-Imam Mohammad Bin Saud Islamic University, Riyadh, Saudi Arabia

**Keywords:** *CYP2C19* genotypes, Pharmacogenetics, clopidogrel, Arab population, platelet reactivity, cardiovascular events

## Abstract

This study aimed to predict the preventive effect of clopidogrel against recurrent cardiovascular events (CVEs) among the Arab population carrying different *CYP2C19* mutations and to determine the frequency of polymorphic alleles and genotypes of *CYP2C19* among them. The review included all the studies that reported data related to the *CYP2C19* polymorphisms among Arab populations. The review included Arab CVDs patients who are categorized into carriers (cases) and non-carriers (controls) of *CYP2C19* alleles and used clopidogrel as secondary prophylaxis. The patients who had recurrent CVEs or had high on-treatment platelet reactivity (HTPR) while using clopidogrel treatment were described as (events). The results showed a significantly increased risk of recurrent CVDs events by about three folds was associated with carriers of *CYP2C19*2* and *CYP2C19*3* mutations compared to non-carriers (OR= 3.32, CI=1.94-5.67, and OR=3.53, CI=1.17-10.63 respectively). However, no significant difference was recorded between both studied groups regarding the presence of *CYP2C19*17* mutation (OR=0.80, (CI=0.44-1.44). The results also revealed that 59 (4.16%) of Arabs carrying two *CYP2C19*2* alleles (homozygous), and 356 (25.12%) have one *CYP2C19*2* allele and one *CYP2C19*1* allele (heterozygous). Moreover, 42 (2.96%) were carrying two *CYP2C19*17* alleles (homozygous), and 262 (18.49%) were carrying one *CYP2C19*17* allele and one wild-type allele of *CYP2C19* (heterozygous). The most common *CYP2C19* genotypes reported among Arabs was the wild-type *\*1/*1*, of which 49.26% of them had the homozygous form of the *CYP2C19*1* allele. The frequency of the CYP2C19*1 allele was 71.07%, followed by the *CYP2C19*2* allele (16.73%) and *CYP2C19*17* (12.21%), respectively. The *CYP2C19*3* allele was detected rarely among Arabs (<1%) compared to *CYP2C19*1, *2*, and *\*17* alleles. The present study revealed that Arabs carrying *CYP2C19*2* and *CYP2C19*3* alleles may not respond to clopidogrel and may put those patients at risk of recurrent CVEs. Carriers of the *CYP2C19*17* allele, on the other hand, did not show a significant role either in increasing or decreasing the antiplatelet efficacy of clopidogrel. The *CYP2C19* genotypes including *\*1/*1, *1/*2, *1/*17, *2/*2*, and *\*17/*17* are commonly distributed among the Arabs.

## 1. Background

Patients with cardiovascular diseases (CVDs) are usually caused by a blood coagulation that blocks the cardiac vessels from supplying oxygen, glucose, minerals, and micro minerals to the cardiac tissues. Therefore, patients with CVDs should be subjected to antiplatelet or anticoagulant medications to avoid further coagulation. Otherwise, they will be at high risk of cardiovascular events (CVEs) in the future [1,2].

Clopidogrel is one of the medications that act as an antiplatelet by inhibiting the binding of adenosine diphosphate (ADP) to P2Y12 receptors, thus avoiding platelets adhesions and aggregations [3].

Clinically, clopidogrel is frequently used in Arab countries as a protective agent to avoid recurrent CVEs. However, several clinics did not have specific criteria to monitor the antiplatelet efficacy of clopidogrel either in the short or long term after the dispensation. Globally, the clinical efficacy of clopidogrel is controvertible, as most of the studies suggest that some genetic mutations among specific ethnicities are described as the causes of weak antiplatelet activity. Therefore, pharmacokinetics and pharmacodynamics studies have been conducted to reveal the basis of this issue.

The pharmacokinetic studies showed that clopidogrel prodrug needs to be activated by several metabolic enzymes. The clopidogrel agent is converted to 2-oxo-clopidogrel (inactive metabolite) by CYP1A2, CYP2C19, and CYP2B6 oxidative enzymes. The 2-oxo-clopidogrel is further converted to cis-thiol-clopidogrel (active metabolite) by CYP3A4, CYP2C19, CYP2C9 and CYP2B6 oxidative enzymes [4,5]. Therefore any mutation of these enzymes, including CYP2C19, could affect the metabolism of clopidogrel and consequently affect its plasma concentration levels and efficacy[6].

More than 30 mutations of *CYP2C19* have been identified [7]; however, only three polymorphic alleles are more familiar, including *CYP2C19*2, CYP2C19*3*, and *CYP2C19*17. CYP2C19*1* is encoding a normal active form of the CYP2C19 enzyme. *CYP2C19*2* and *CYP2C19*3* mutant alleles encode an inactive CYP2C19 enzyme, and the *CYP2C19*17* mutant allele is known to express a more active form of CYP2C19 enzyme [8].

Previous studies revealed that mutations of *CYP2C19* alleles are highly distributed among Asian people but lower among Europeans [9,10]. Nevertheless, the mutations of these alleles are not well studied among the Arab population. Therefore, this study aimed to investigate the efficacy of clopidogrel as an antiplatelet drug among those carrying these alleles and genotypes and determine the frequency of *CYP2C19* polymorphic alleles and genotypes among the Arab population.

## 2. Methods

### 2.1 Search methods

Two authors independently searched in PubMed, Google Scholar, and EMBASE databases for published English studies at any year related to clopidogrel efficacy and the frequency of *CYP2C19* gene polymorphism among Arabs. The exclusion criteria comprised non-Arab populations and patients with any contraindication for clopidogrel. The following terms were used in the search; *CYP2C19* Genotypes; OR *CYP2C19* Polymorphic alleles; OR *CYP2C19* Gene Mutations; AND clopidogrel response; OR Antiplatelet activity; AND Arabs.

### 2.3 Type of participants

This systematic review (for the qualitative part) included Arab people who were genotyped to determine the frequency of *CYP2C19* genotypes and alleles. Concerning the quantitative part (meta-analysis), only patients who previously had CVEs and using clopidogrel as secondary prophylaxis had been included. Those patients were categorized into carriers of (cases) and non-carriers (controls) of *CYP2C19* mutations, including *CYP2C19*2, CYP2C19*3*, and *CYP2C19*17*.

### 2.3 Outcome measures

The outcome of the quantitative part was to predict the antiplatelet efficacy of clopidogrel among carriers and non-carriers of *CYP2C19* mutations who previously had CVEs through measuring the frequency of the recurrent CVDs or high platelet aggregation reported. The outcome of the qualitative part was to determine the frequency of genotypes and alleles related to the *CYP2C19* gene among Arabs.

### 2.4 Data collection and extraction

Two authors independently reviewed the abstracts of potential articles for inclusion criteria and obtained all the relevant articles. Then, they extracted the following characteristics from the included studies; study setting, duration, design, participants’ age, sex, and outcome measures. Disagreement was solved by discussion.

### 2.5 Assessment of the risk of bias, quality of evidence, and treatment effect

Two authors independently assessed the risk of bias of the included studies. They graded each risk of bias as high, low, or unclear according to The Newcastle Ottawa Scale for non-randomized Studies [11]. According to the GRADE approach, the quality of evidence for each outcome measure was judged as high, moderate, low, or very low (Grading of Recommendations Assessment, Development, and Evaluation) [12]. The researchers analyzed the data using the Review Manager 5.3 program [13]. The risk of recurrent CVDS events was measured using Odds Ratio (OR) with 95% Confidence Interval (CI).

### 2.6 Dealing with heterogeneity

The I2 statistic was used to assess heterogeneity among the included studies in each analysis [14].

## 3. Results

### 3.1 Results of Search

One hundred and eighty-eight potentially relevant articles were searched, 97 were identified after the removal of duplicates. Two authors independently reviewed the abstracts of the articles based on inclusion and exclusion criteria. Nineteen full-text articles were assessed for eligibility; ten of them met the inclusion criteria (10 in qualitative and 6 in quantitative analysis). Details of the search are given in the PRISMA flow diagram (see figure.1)

**Figure.1.**
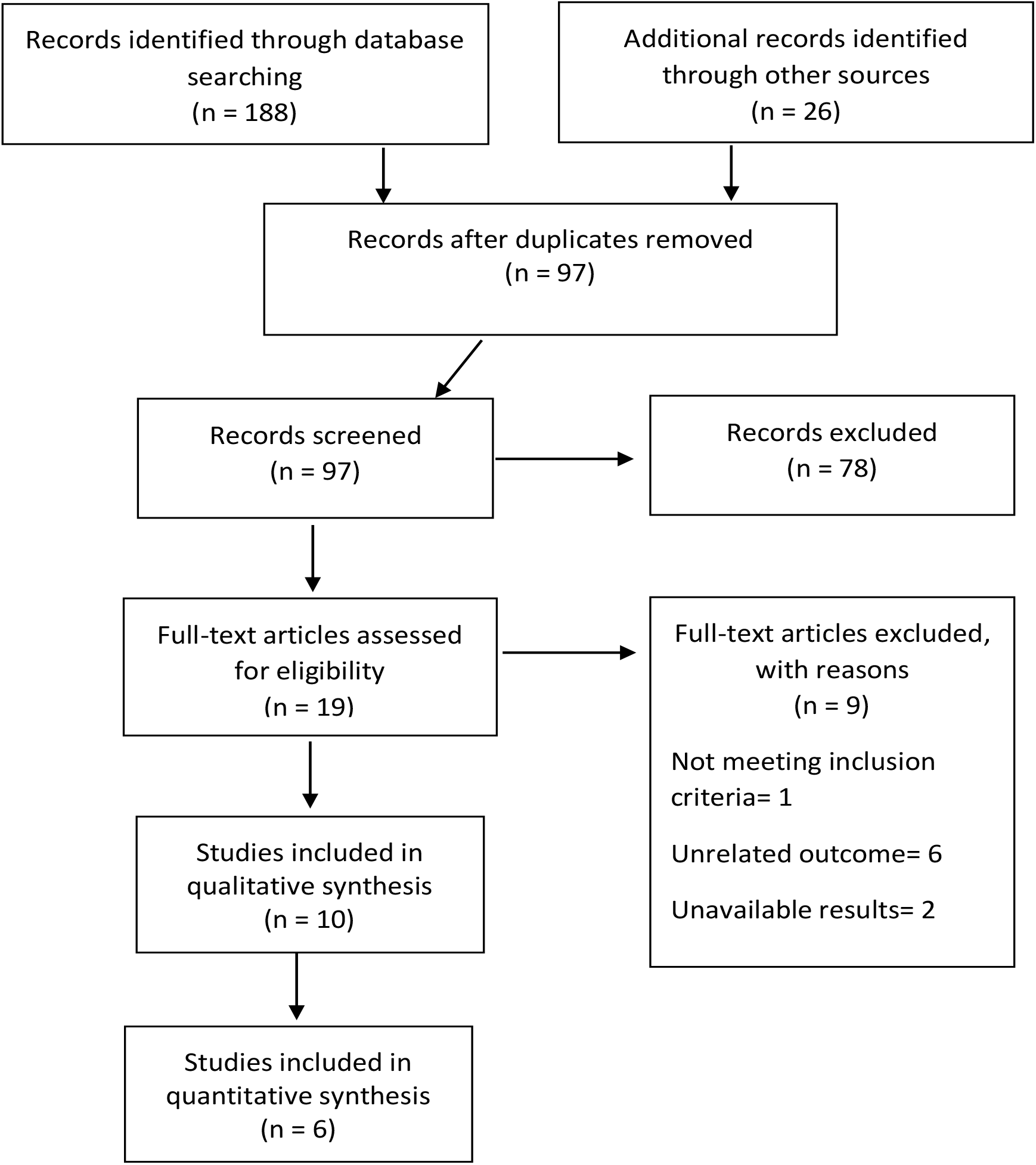
PRISMA flow diagram.

### 3.2 Included Studies

In the quantitative part, the review included 6 observational studies (2 retrospectives, 3 prospective, and one cross-section) [8,15–19], which reported data for clopidogrel antiplatelet efficacy in the presence of different *CYP2C19* gene variations.

On the other hand, the qualitative part included 10 studies, four case series, and six observational studies. These ten studies reported the frequency of *CYP2C19* polymorphisms among Arabs [20–23].

Two authors independently extracted characteristics of the included studies, including study title, journal, study design, duration, setting, aim, participants’ age, sex, number, and outcome measures (see table.1).

**Table.1.** Characteristics of the included studies.

| Author | ID | Setting/duration | Design | Participants | Aim | Outcome |
| --- | --- | --- | --- | --- | --- | --- |
| 1- Adel Alhazzani et al 2017 | Pharmacogenetics of *CYP2C19* genetic polymorphism on clopidogrel response in patients with ischemic stroke from Saudi Arabia | Neurology Clinics at Aseer Central Hospital, Abha, Kingdom of Saudi Arabia, between October 2015 and January 2016 | Retrospective study | 50 patients on 75 mg maintenance dose of clopidogrel therapy suffering from stroke | To assess the influence of *CYP2C19* genetic polymorphisms on the response to clopidogrel in ischemic stroke in Saudi Arabian population. | The variant allele (homozygous and homozygous Mutant) showed significant influence on platelet inhibition and the antiplatelet effect of clopidogrel in ischemic stroke. |
| 2-Khalil et al, 2016 | Genetic and Nongenetic Factors Affecting Clopidogrel Response in the Egyptian Population | Hospital network in Chicago, Illinois between 5 March and 6 April 2020. | Retrospective study | 190 patients with acute coronary syndrome (ACS) treated with clopidogrel (75 mg/day) for at least a month. | To investigate genetic and non-genetic factors associated with clopidogrel response in Egyptians. | *CYP2C19* loss-of-function (LOF) alleles carriers had increased risk of recurrent CVES vs. noncarriers (odds ratio 2.52; 95% confidence interval 1.23–5.15, P = 0.011). |
| 3-Abid et al, 2013 | Impact of cytochrome P450 2C19*2 polymorphism on the clinical cardiovascular events after stent implantation in patients receiving clopidogrel of a southern Tunisian region | Department of Cardiology of Sfax (Tunisia) May 2009 and September 2010 | Prospective study | 100 consecutive patients admitted to the cardiology department for percutaneous coronary stenting. 2 groups: those with at least one *CYP2C19* *2 allele (*2 carriers) and non-carriers. | to investigate the genetic variant of the gene CYP 2C19 in our population To assess the involvement of this genetic profile in the occurrence of major cardiovascular events | The prevalence of *CYP2C19* *2 allele was 11.5%. No statistically significant differences were noted between the two groups regarding the occurrence of intra hospital recurrent CVES. |
| 4- Mohammad et al 2018 | *CYP2C19* genotype is an independent predictor of Adverse Cardiovascular Outcome in Iraqi Patients on Clopidogrel Post Percutaneous Coronary Intervention. | Duhok Heart center, Kurdistan-Iraq, in the period between Jan 2014 to Mar 2017 | Prospective study | 201 unselected patients undergoing percutaneous coronary intervention (PCI) aged 35-82 (M:F=1.9:1) | To determine the impact of *CYP2C19* genotyping on the occurrence of major adverse cardiovascular events (recurrent CVES) | The risk of recurrent CVES is 8.6% after a median follow-up of 12 months, |
|  |  |  |  | 186 patients had regular follow up |  |  |
| 5- Ayesh et al 2017 | The clinical effects of *CYP2C19* *2 allele frequency on Palestinian patients receiving clopidogrel after percutaneous coronary intervention | The cardiology department of the European Gaza Hospital (EGH). March and the end of May 2014 | Prospective study | 110 consecutive unrelated post-PCI patients | To determine the prevalence of *CYP2C19* *2 and *3 alleles in Palestinian patients with percutaneous coronary intervention | The frequency of *CYP2C19* *1, *2 and *3 alleles was 82.3%, 15.5% and 2.3% respectively |
| 6- Al-Azzam et al, 2013 | Factors that contribute to clopidogrel resistance in cardiovascular disease patients: Environmental and genetic approach | King Abdullah University Hospital (KAUH) and Jordan University Hospital (JUH) in the period between November 2010 and June 2011. | Cross-section study | 270 cardiovascular disease patients | To investigate factors that contribute to clopidogrel resistance | Gender, concomitant use of calcium channel blockers, HDL and *CYP2C19* *2 polymorphism are significant factors |
| 7- Ahmad et al 2018 | Analysis of Gene Polymorphism *CYP2C19* in the Lebanese Population Who Reside in Colombia | three Lebanese volunteers residents of Colombia | Cross-section study | 109, 38 women and 71 men between 18 and 75 years | To determine the polymorphism of the *CYP2C19* gene in the Lebanese | similar behavior with the alleles frequencies of the previous studies made in Colombia, Africa, Europe and other American population. |
| 8-Rjoub et al 2018 | Allelic frequency of *PON1 Q192R*, *CYP2C19* *2 and *CYP2C19* *17 among Jordanian patients taking clopidogrel | among Jordanian patients in University of Jordan | Cross-section study | 148 unrelated Jordanian patients who were taking clopidogrel (55 females and 95 males) | To investigate the influence of allelic frequencies of *PON1 Q192R*, *CYP2C19* *2 and *CYP2C19* *17 genetic polymorphisms on the response to clopidogrel | The *CYP2C19* *2, *CYP2C19* *17, and *PON1 Q192R* allele frequencies were 9.8, 28.72 and 28.7 %, respectively |
| 9- Khalaf et al 2016 | Impact of *Cytochrome P450 2C19* *2 and *3 on Clopidogrel Loading Dose in Saudi Patients with Acute Coronary Syndrome | Prince Sultan Cardiac Center, Buraidah, Saudi Arabia | Prospective study | 90 patients underwent coronary angioplasty with drug eluting stents. | To evaluate the impact of *CYP2C19* allele *2 and allele *3 on PRU and the potential clinical consequences of such interaction. | Genotypes *1/*1, *2/*2, and *1/*2 were expressed by 60, 28, and two patients (67, 32, and 3%), respectively |
| 10- Al-Jenoobi et al 2012 | *CYP2C19* Genetic Polymorphism in Saudi Arabians | King Saud University, Riyadh, Saudi Arabia Different | Cross-section study | 192 healthy unrelated Saudi Arabians | To evaluate *CYP2C19* genetic polymorphism in a Saudi Arabian population by determining frequencies of | The allelic frequency of heterozygous *CYP2C19* *2 was 8.2% with only one individual |
|  |  | geographic regions |  |  | *CYP2C19* *2, *3, *4, *6, *7 and *17 alleles | carry the homozygous genotype of this defective allele |

### 3.3 Trial Participants

The quantitative part of the review included 878 Arabic (Saudis, Egyptians, Jordanians, Tunisians, Iraqis, and Palestinians) patients who were previously diagnosed with CVDs. For the qualitative part, 1417 Arabic people, either healthy or non-healthy, were included. The participants in the qualitative part originated from 7 different Arabic countries, including Saudi Arabia, Egypt, Jordan, Lebanon, Tunisia, Iraq, and Palestine.

### 3.4 Risk of bias among included Studies

Overall, no high risk of bias was recorded among the included studies in this review. Regarding {adequate case definition} bias and {comparability of cases and control} bias, it was unclear in Rjoub et al. study. Also, the {same representative rate for cases and control} bias was low risk in all included studies. While {consecutive representative of cases} and {independent outcome assessment} bias, it was unclear in most of the included studies (see figure.2).

**Figure.2.**
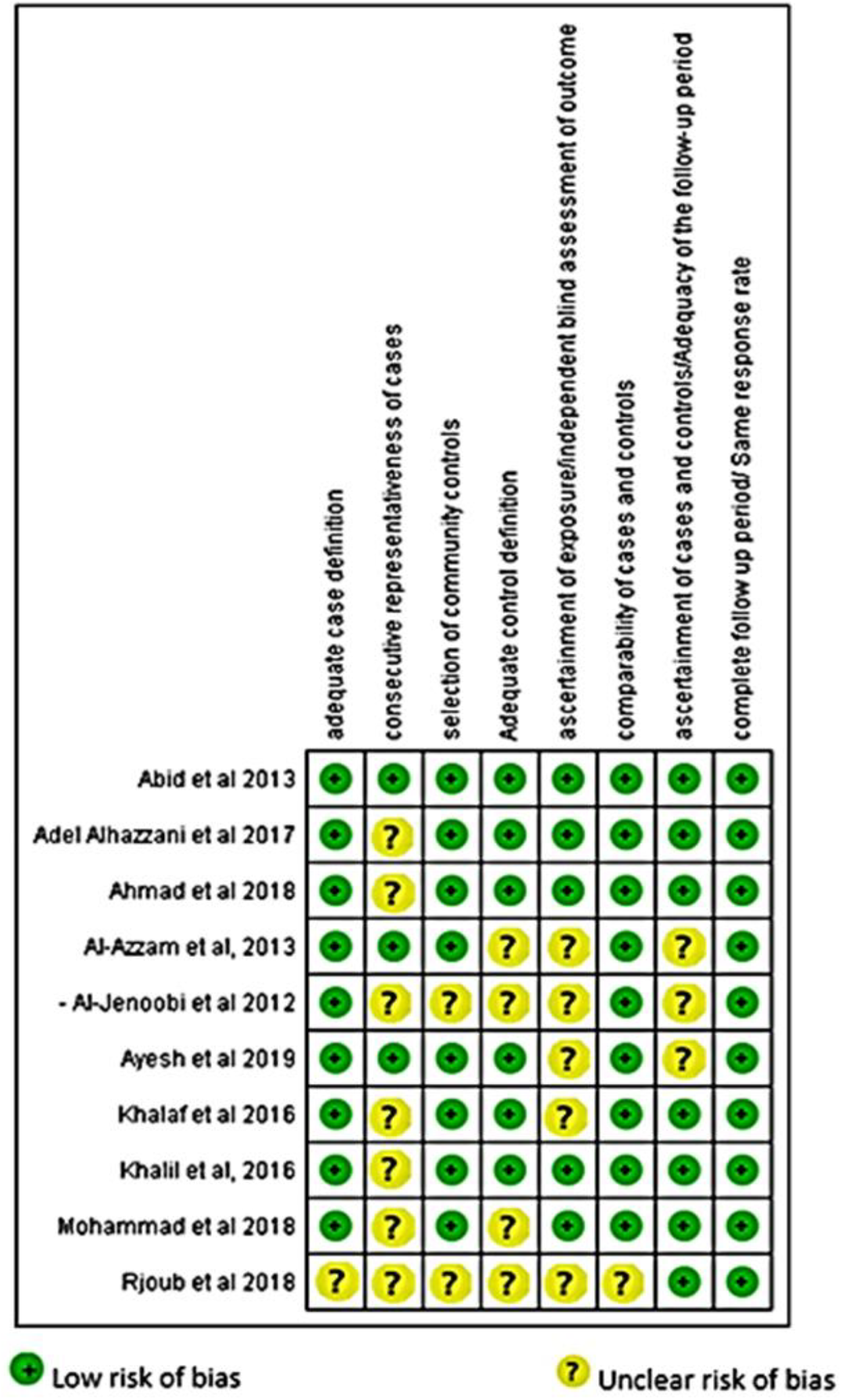
Risk of bias among included studies.

### 3.5 Outcomes

#### 3.5.1 Outcomes of the quantitative part

Figures 3,4 and 5 presents the forest plots of the frequency of recurrent CVDs events among Arabic patients using clopidogrel either carrying or not carrying *CYP2C19*2, CYP2C19*3*, and *CYP2C19*17* mutations. The results showed a significantly increased risk of recurrent CVDs events by about three folds was associated with carriers of *CYP2C19*2* and *CYP2C19*3* mutations compared to non-carriers (OR= 3.32, CI=1.94-5.67, and OR=3.53, CI=1.17-10.63 respectively). However, no significant difference was recorded between both studied groups regarding the presence of *CYP2C19*17* mutation (OR=0.80, (CI=0.44-1.44).

**Figure.3.**
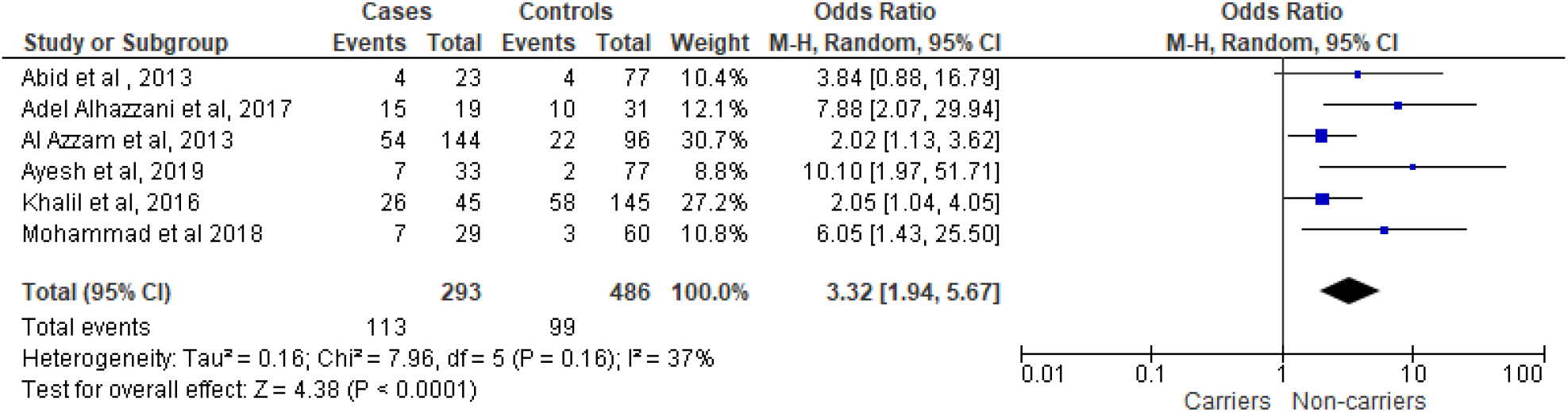
Forest plot of the frequency of recurrent CVDs among carriers and non-carriers of *CYP2C19*2* mutation.

**Figure.4.**
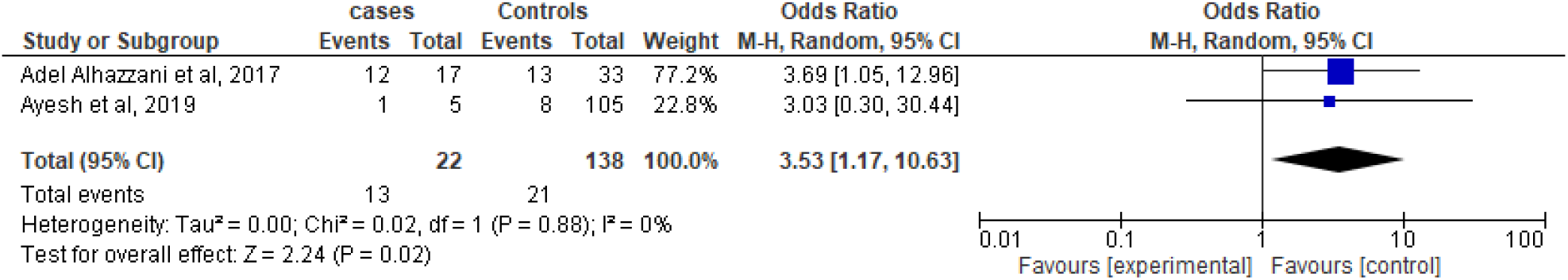
Forest plot of the frequency of recurrent CVDs among carriers and non-carriers of *CYP2C19*3* mutation.

**Figure.5.**
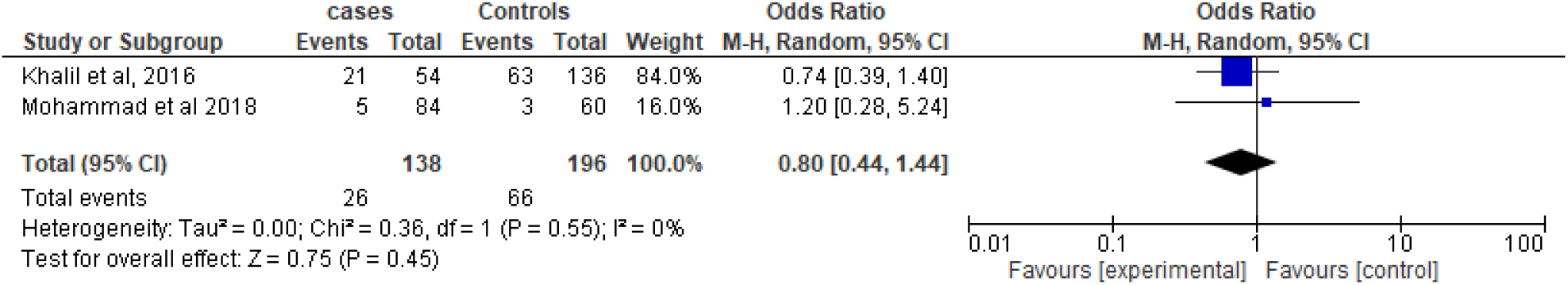
Forest plot of the frequency of recurrent CVDs among carriers and non-carriers of *CYP2C19*17* mutation.

#### 3.5.2 Outcomes of the qualitative part

This part included 1417 Arabic people, which were genotyped in order to determine the *CYP2C19* gene variations and to detect the availability of any well-known mutated alleles, including *CYP2C19*2, CYP2C19*3*, and *CYP2C19*17* alleles among Arab populations, including Saudis, Egyptians, Jordanians, Iraqis, Palestinians, Lebanese and Tunisians people. The results revealed that 59 (4.16%) of them were carrying two *CYP2C19*2* alleles (homozygous), and 356 (25.12%) had one *CYP2C19*2* allele and one *CYP2C19*1* allele (heterozygous). Moreover, 42 (2.96%) were carrying two *CYP2C19*17* alleles (homozygous), and 262 (18.49%) were carrying one *CYP2C19*17* allele and one wild-type allele of

##### *CYP2C19* gene (heterozygous)

The most common CYP2C19 genotype reported among Arabs was the wild-type *\*1/*1*, of which 49.26% of them had the homozygous form of the *CYP2C19*1* allele. The frequency of the *CYP2C19*1* allele was 71.07%, followed by the *CYP2C19*2* allele (16.73%) and *CYP2C19*17* (12.21%), respectively. The *CYP2C19*3* allele was detected rarely among Arabs (<1%) compared to *CYP2C19*1, *2*, and *\*17* alleles.

Based on the frequencies of genotypes, about half of Arab people (>49%) could be described as CYP2C19 extensive metabolizers. Other common *CYP2C19* gene phenotypes that existed among Arabs are intermediate metabolizers (25%), rapid metabolizers (18%), poor metabolizers (4%), and ultra-rapid metabolizers (3%), respectively.

## 4. Discussion

Many studies genotyped the *CYP2C19* gene to assess clopidogrel’s efficacy among specific ethnic groups. However, few studies correlated *CYP2C19* gene mutation and clopidogrel efficacy among Arab ethnic groups.

In the quantitative part, the present study recorded significant differences between carriers (cases) and non-carriers (controls) of *CYP2C19*2, *3* alleles regarding the number of recurrent CVEs in Arabs using clopidogrel (OR=3.32, CI=1.94, 5.67, and OR=3.53, CI=1.17, 10.63, respectively). However, there was no statistical difference among carriers and non-carriers of *CYP2C19*17* allele concerning the same aspect (OR=0.80, CI=0.44, 1.44).

These results indicate that Arab patients carrying *CYP2C19*2* and *\*3* alleles may decrease the antiplatelet activity of clopidogrel and could lead to recurrent CVEs. The present outcomes were consistent with more than 18 high-quality clinical trials and 6 meta-analysis studies. They revealed that *CYP2C19*2* and *\*3* alleles have a significant role in causing recurrent CVEs among patients using clopidogrel [24–43].

On the other hand, 3 previous meta-analysis studies concluded that loss-of-function alleles (*CYP2C19*2* and *CYP2C19*3*) have no significant effect in causing recurrent CVEs while using clopidogrel. However, they showed a significant effect in leading to other complications (e.g., ST-elevation and stent thrombosis) [44–46]. This could be explained by the presence of other genetic factors that may affect both the clopidogrel bioactivation process and recurrent CVEs, including specific *CYP2C9, CYP3A4, CYP1A2*, and *CYP2B6* genes’ mutations.

Concerning the qualitative part, the study revealed that CYP2C19 genotypes including *\*1/*1, *1/*2, *1/*17, *2/*2*, and *\*17/*17*, respectively, are commonly distributed among Arabs. Compared with other ethnic groups, the *CYP2C19*1* allele among Arabs was more or less similar to Caucasians (59.2%), Africans (70.2%), and Asians (65%). Also, the *CYP2C19*2* allele is similar to that of Caucasians (15.1%) and Africans (12.6%), but less than Asians (34.5%). The *CYP2C19*3* allele was rare among all ethnic groups, except Asians (9%). While the *CYP2C19*17* allele represented 12.2% among Arabs, it was recorded as the highest frequent mutant allele among the Caucasian population (25.7%), followed by the Africans (17.2%) and Asians (0.5%) [24].

## 5. Conclusions

The present study revealed that Arabs carrying *CYP2C19*2* and *CYP2C19*3* alleles may not respond to clopidogrel and may put those patients at risk of recurrent CVEs. Carriers of the *CYP2C19*17* allele, on the other hand, did not show a significant role either in the increase or decrease of the antiplatelet efficacy of clopidogrel. The *CYP2C19* genotypes including *\*1/*1, *1/*2, *1/*17, *2/*2*, and *\*17/*17* are commonly distributed among the Arabs.

## Data Availability

All data produced in the present study are available upon reasonable request to the authors

## Declarations

### Availability of data and material

The authors confirm that the data supporting the findings of this study are available within the article.

## Acknowledgments

The researchers would like to thank Dr. Yousef Almutairi (Saudi Ministry of Health, Riyadh, Saudi Arabia) for their assistance in reviewing the manuscript.

## Author Contributions

AK and NR contributed in conceptualization. AK, NR and NM contributed in writing – original draft preparation. NM, AF, and KA contributed in writing – review and editing. NR, NM, AK, and AF contributed in resources. All authors have read and agreed to the published version of the manuscript.

## Ethics approval and consent to participate

Not applicable.

## Consent for publication

Not applicable.

